# Leveraging genetics to improve Body Mass Index increase prediction in the first-episode of psychosis

**DOI:** 10.1101/2022.02.15.22270982

**Authors:** Gerard Muntané, Javier Vázquez-Bourgon, Ester Sada, Lourdes Martorell, Sergi Papiol, Elena Bosch, Arcadi Navarro, Benedicto Crespo-Facorro, Elisabet Vilella

**Author notes:** Corresponding authors: Gerard Muntané; Javier Vázquez-Bourgon. These authors share senior authorship (Benedicto Crespo-Facorro, Elisabet Vilella).

## Abstract

**Background:** Individuals with a first episode of psychosis (FEP) show rapid weight gain during the first months of treatment, which is associated with reduction in psychiatric and physical health. Although genetics is assumed to be a significant contributor to this weight gain, its exact role is unknown.

**Methods:** Reference GWAS from BMI and SCZ were obtained to evaluate the pleiotropic landscape between both traits using the pleioFDR software. In parallel, we gathered a population-based FEP cohort of 381 individuals and BMI Polygenic risk-scores (PRS) were evaluated on a sample of 224 individuals. Subsequently, the PRSs obtained from both BMI and the variants shared between the two traits were incorporated into risk models that included demographic and clinical variable to predict BMI increase (ΔBMI) on an independent sample of 157 patients.

**Results:** BMI PRS significantly improved the prediction of absolute BMI and ΔBMI during the first 12 months after the onset of psychotic symptoms. This improvement, was mainly explained by shared variants between SCZ and BMI. In contrast, absolute BMI was predicted mainly by non-shared variants.

**Conclusions:** We validated, for the first time, that genetic factors play a key in the determination of both BMI and ΔBMI in FEP. This finding has important clinical implications in identifying individuals who require specific treatment strategies. Improved risk classification may help prevent associated adverse metabolic events, and reduce overtreatment and costs for both individuals and the healthcare system. It also highlights the importance of studying genetic pleiotropy in the context of medically important comorbidities.

## Introduction

Schizophrenia (SCZ) is a severe disorder with a high burden of morbidity and societal impact. It has a heritability of ∼80%, much of which is attributable to common risk alleles (1). Patients with SCZ present with a high number of comorbid medical conditions across their lifespan (2,3). In fact, people with SCZ have a higher mortality rate than the general population, corresponding to a 10-20 year reduction in life expectancy, predominantly, due to cardiovascular disease (4,5).

Antipsychotic (AP) drugs can help reduce the intensity and frequency of psychotic symptoms, but on the other hand, they are obesogenic (6). Obesity represents a modifiable risk factor related to adverse health-related outcomes (7), reducing quality of life (8) and adherence to treatment (9). The risk of obesity in patients with SCZ is more than four times higher than in the general population (10). The first year after the initiation of AP treatment is a critical period, in which up to 60-80% of the total weight gain occurs (11,12). Among all factors studied, rapid initial weight gain, AP drug, pre-treatment body mass index (BMI), and sex are the best predictors of weight gain and associated metabolic abnormalities (13–15). However, substantial differences in susceptibility between individuals suggest that weight gain may be partially explained by a mixture of environmental effects and genetic background (16–18). In support of this view, the heritability of weight gain in monozygotic twins has been estimated to be 0.6-0.8 and a few genetic loci have been associated with AP-induced weight gain by genome-wide association studies (GWAS) (19–21). Incidentally, SCZ, given their strong polygenic component, exhibit extensive genetic overlap with other mental disorders (22,23), but also with other non-psychiatric traits (24–26). In particular, studies reported a negative overall genetic correlation between BMI and SCZ, resulting from a mixture of variants with positive and negative effects (26–28).

There is substantial genetic vulnerability to BMI trajectories in the general population (29,30); but it remains to be determined whether the Polygenic Risk Score (PRS) for BMI has any clinical utility in populations with psychiatric disorders. However, the extensive genetic overlap between BMI and SCZ suggests the possibility of developing prediction models including genetic factors. In this study, we aimed to find out whether common genetic variants confer a risk for increased BMI in FEP patients, and to investigate the role of shared variants between SCZ and BMI in such BMI increase (ΔBMI). To our knowledge, this is the first study to demonstrate the importance of genetics in FEP-associated ΔBMI which has the potential to identify individuals at increased risk at the early stages of the disease.

## Methods and Materials

### Study design

This study includes individuals with a FEP initially enrolled in the Cantabria Program for Early Intervention in Psychosis (PAFIP program, Spain) between 2001 and 2018 (31). Inclusion criteria were: age 16-60 years; living in the catchment area; experiencing their first episode of psychosis (FEP); no prior treatment with AP medication or, if previously treated, a total life-time of adequate AP treatment of less than 6 weeks. The diagnoses were confirmed using the Structured Clinical Interview for DSM-IV1 (SCID–I) conducted by an experienced psychiatrist at six months from the baseline visit. DSM-IV criteria for drug or alcohol dependence, intellectual disability and having a history of neurological disease or head injury were exclusion criteria. Written informed consent was obtained from all subjects. The Clinical Research Ethics Committee of Cantabria approved the research protocol.

Patients were assigned in 3 consecutive phases of the PAFIP (PAFIP I, II and III), encompassing randomized, flexible-dose, and open-label clinical trials. Patients were randomly assigned to treatment and followed-up for 12 months. During this period, AP doses were adjusted at the treating psychiatrist’s discretion, in an attempt to target the lowest effective dose (32). AP polypharmacy, although not recommended, was allowed if clinically justified. BMI of patients was assessed at baseline (BMI_0_) and re-assessed after 3 (BMI_3_) and 12 months (BMI_12_) of treatment initiation. Together with BMI, AP drug prescribed, dose of chlorpromazine equivalents, and whether patients were on antidepressant treatment were collected at every time point. Patients were grouped based on the primary active drug and doses of chlorpromazine equivalents were calculated (33).

### Samples and Genotyping

The PAFIP sample was divided into two independent patient datasets. The first dataset including 224 patients was used for discovery (from now on *Training set*), while a second sample including 157 patients was used for replication (from now on *Validation set*). In both datasets, DNA was extracted from peripheral lymphocytes, and genotyping was performed using the Affymetrix 6.0 platform (*Training set*) and the Illumina Infinium PsychArray (*Validation set*), respectively. Standard quality-control procedures (34) were performed with PLINK 1.9 (35), resulting in 6,910,431 SNPs in the *Training set* and 6,552,380 SNPs in the *Validation set* (Supplemental Information). Finally, only those individuals with valid BMI measures were included (Table S1).

### Pleiotropy analyses from GWAS data

GWAS summary statistics on BMI were obtained (36), which comprised association analyses of a total of 806,834 European individuals. For SCZ, we obtained GWAS data on 67,390 patients and 94,015 controls, with 80% being of European ancestry (1) (Table S2). GWAS summary statistics were referenced to 9,546,816 SNPs generated from the 1,000 Genomes Project (1KGP, https://www.internationalgenome.org/). SNPs that were non-biallelic, without rsIDs, duplicated, or with strand-ambiguous alleles were removed. We also filtered out SNPs with INFO scores⍰<⍰0.9, those mapping to the extended major histocompatibility complex (MHC, chr6: 25,119,106–33,854,733), SNPs located on chromosomes X, Y and mitochondria, and SNPs with sample sizes 5 standard deviations away from the mean. Finally, a common set of 1,949,409 SNPs were kept in both datasets. ORs and betas were transformed into z-scores.

We used *pleioFDR* (https://github.com/precimed/pleiofdr) to identify genetic loci jointly associated with the two phenotypes, as previously described (25) (Supplemental information). SNPs jointly associated with BMI and SCZ (conjFDR<0.05) were mapped to genes with ANNOVAR(37). Genelists were submitted to KOBAS-I (38) to check for enrichment in biological categories and diseases; and to the GENE2FUNC option implemented in FUMA to check for tissue-enrichment (39). All protein-coding genes were used as background and the Benjamini-Hochberg (BH) method was used for false discovery rate (FDR) correction.

To obtain the widest representation of the SNPs in loci shared between SCZ and BMI, SNPs that were in r^2^>0.1, distance<250kb, and MAF>0.001 with each independent SNPs at conjFDR<0.05 were recovered with PLINK v1.9 from the CEU population of the 1kG project. After this process, all the obtained SNPs were considered as pleiotropic SNPs. Non-pleiotropic SNPs were defined as all the SNPs in the original BMI GWAS that were not contained in any pleiotropic locus.

### Polygenic Risk Scores estimation

A Polygenic Risk Score (PRS) is a measure of an individual’s genetic load of risk alleles for a certain trait. PRSice 2.3.1.e was used to implement a pipeline in PRS creation (40). The GWAS on BMI (36) was used as base data to calculate PRSs in our sample. We performed P-value-informed clumping with PLINK using a cutoff of r^2^⍰=⍰0.1, 250-kb window, SNPs with INFO>0.9, and excluding the MHC. Then, we calculated scores for multiple p-thresholds (from 5e-08 to 1 with increments of 5e-05).

To calculate PRS we used a two-step procedure as previously developed (41), avoiding sample overlap between the *Training* and *Validation sets* that prevents test statistic inflation. First, PRSs were calibrated in the *Training set* and the optimal p-value (P_optimal_) threshold was determined as the one with the highest prediction for each phenotype (BMI and ΔBMI at baseline, 3, and 12 months). Sex, age and the 10 first PCs were included as covariates (when evaluating ΔBMIs, BMI_0_ was also included as covariate). Then, SNPs that fell below each obtained P_optimal_ threshold were identified and carried forward for PRS computation in individuals of the *Validation set*.

### Statistical Analyses

To elucidate which variables were associated with BMI and ΔBMI in our dataset we carried out multiple linear regression followed by type II ANOVA, to test the improvement in the predictive ability of the PRS inclusion. The linear models included the 10 first PCs (to adjust for population stratification), sex, age, AP dose and the type of AP drug (at 3 and 12 months), the diagnosis, and use of antidepressants. Whenever necessary, ΔBMI_3_ and BMI_0_ were also included. These models combining only clinical and demographic variables were referred to as *Null models*. Then, to examine the predictive ability of BMI PRS, we created a series of absolute risk models including genetic factors, that consisted of multiple linear regression analyses with the observed BMI (or ΔBMI) as the dependent variable and the covariates in the *Null model* plus the P_optimal_ PRS, named as *PRS models*. To decipher the role of variants jointly associated with BMI and SCZ, we independently evaluated the *PRS models* using PRS derived from both pleiotropic and non-pleiotropic loci. In those cases, only SNPs from each category were included. Finally, Kruskal-Wallis test was used to compare whether the AP drug prescribed affected ΔBMI at 3 months (ΔBMI_3_) and ΔBMI at 12 months (ΔBMI_12_). All the analyses were performed in R 3.6.0 (42).

## Results

### Characteristics of the sample

A total of 381 patients were included. Among them, 188 (49.3%) were diagnosed with schizophrenia, 115 (30.2%) with schizophreniform disorder, 43 (11.3%) with brief psychotic disorder, 28 (7.4%) with unspecified psychosis, 5 (1.3%) with schizoaffective disorder, and 2 (0.5%) with delusional disorder. At the end of the study, 89.2% of the patients had all BMI measures (Table S1). None of the participants was initially treated with clozapine, which is indicated in treatment-resistant SCZ (43) (Table 1).

**Table 1.** Number of participants in each treatment category at baseline and after 3 and 12 months of follow-up.

| *drug* | *t=0* |  | *t=3months* |  | *t=12 months* |  |
| --- | --- | --- | --- | --- | --- | --- |
|  | *Training* | *Validation* | *Training* | *Validation* | *Training* | *Validation* |
| Aripiprazole | 101 (45.1%) | 11 (7%) | 81 (38%) | 9 (5.8%) | 81 (38.6%) | 9 (6.3%) |
| Clozapine | - | - | 3 (1.4%) | - | 10 (4.8%) | 2 (1.4%) |
| Trifluoperazine | - | - | 1 (0.5%) | - | - | - |
| Haloperidol | - | 34 (21.7%) | - | 31 (20.1%) | - | 17 (11.9%) |
| Olanzapine | 1 (0.4%) | 42 (26.8%) | 13 (6.1%) | 45 (29.2%) | 24 (11.4%) | 46 (32.2%) |
| Paliperidone | - | - | 3 (1.4%) | - | 12 (5.7%) | - |
| Quetiapine | 34 (21.7%) | 15 (9.6%) | 14 (6.6%) | 12 (7.8%) | 13 (6.2%) | 18 (12.6%) |
| Risperidone | 49 (21.9%) | 43 (27.4%) | 71 (33.3%) | 49 (31.8%) | 54 (25.7%) | 44 (30.8%) |
| Ziprasidone | 39 (24.8%) | 12 (7.6%) | 27 (12.7%) | 8 (5.2%) | 15 (7.1%) | 7 (4.9%) |
| Amisulpride | - | - | - | - | 1 (0.5%) | - |
| **TOTAL** | **224** | **157** | **213** | **154** | **210** | **143** |

In the whole dataset, the mean ΔBMI_3_ and ΔBMI_12_ were 6.4% and 13.9%, respectively (Figure S1). Most of the individuals displayed a positive ΔBMI_12_ (n=312, 91.8%), 25 individuals (7.4%) showed a negative ΔBMI_12_, and 3 individuals (<0.1%) presented the same BMI_0_ and BMI_12_. In total, 108 patients in the *Training set* (48.2%) and 94 in the *Validation set* (59.9%) remained with the same AP drug during the study (Figure S2). Both ΔBMI_3_ and ΔBMI_12_ were negatively correlated with BMI_0_ (P=5.56e-10 and P=7.07e-07) and positively correlated among them (Figure S3). All APs were associated, on average, with a positive ΔBMI (Figure S4 and Supplemental Information).

### Pleiotropy between SCZ and BMI

A total of 486 independent SNPs belonging to 449 near-independent genomic loci (r^2^⍰<⍰0.1) were identified to be jointly associated with SCZ and BMI at conjFDR⍰<⍰0.05 (Figure 1A and Table S3). Among them, 169 independent SNPs (35%) showed agonistic relationships between SCZ and BMI, where the alleles that confer risk for SCZ, also increase BMI. In addition, 317 independent SNPs (65%) showed antagonistic links, where the allele that increased SCZ-risk, had a BMI reduction effect (Figure 1B). This represented an approximate two-fold ratio between negative (antagonistic) and positive (agonistic) pleiotropic variants (Figure S5). Both groups of variants did not differ in their effects in SCZ (P=0.45) or BMI (P=0.17), nor in their minor allele frequency (P=0.72).

**Figure 1.**
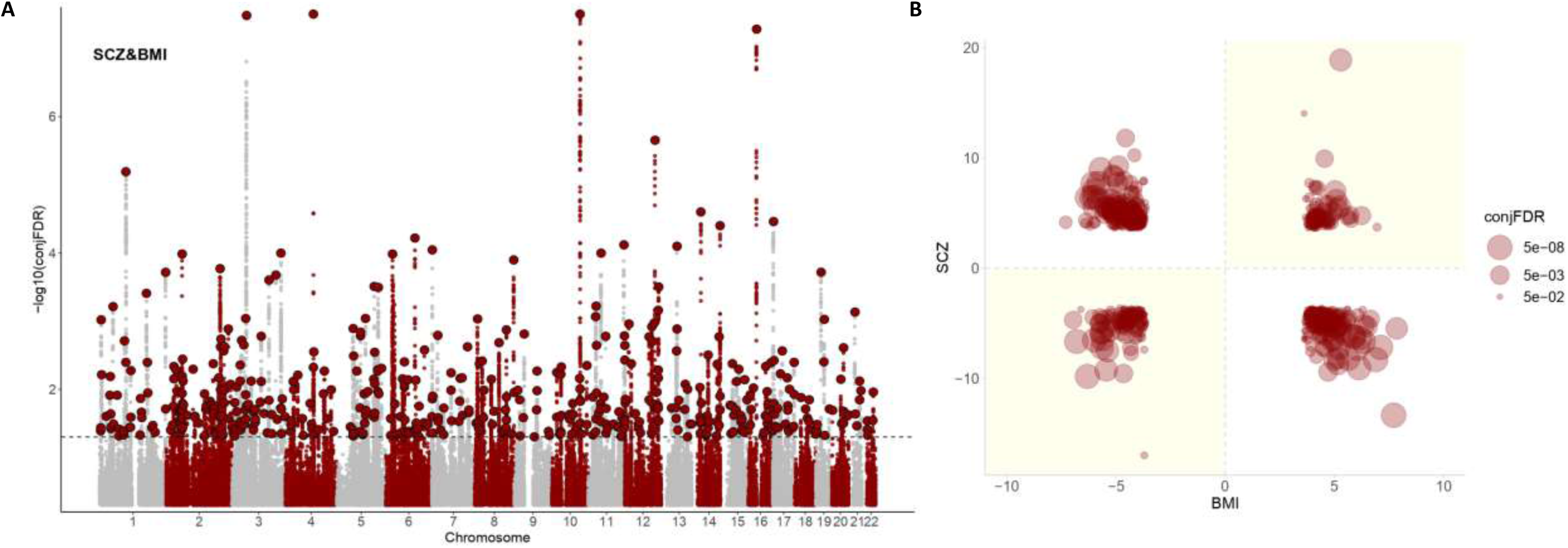
Pleiotropic variants between SCZ and BMI. A) Manhattan plot showing independent (r^2⍰^<⍰0.1) loci associated with both SCZ and BMI, as defined by conjunction false discovery rates (conjFDR) after excluding SNPs in the MHC region. The dashed black line represents the conjFDR threshold of 0.05. B) Pleiotropy plot for independent SNPs with conjFDR⍰<⍰0.05 (n⍰=⍰486) between SCZ and BMI. The conjFDR values and the direction of the effects (z-scores) of the minor alleles are plotted for BMI (x-axis) against SCZ (y-axis). Graph regions whose effects are consistent with a positive correlation between both traits are shadowed in yellow. C) Ratio between negative and positive pleiotropic variants across different conjFDR thresholds.

The whole set of SNPs shared between SCZ and BMI included 9,262 SNPs corresponding to 889 genes in total, which were enriched in DNA methylation (FDR=4,99e-14), DNA damage (FDR=7,52e-13), cAMP signaling pathway (FDR=2,45e-09), axon guidance (FDR=1,06e-07), dopaminergic synapse (FDR=2,99e-05), insulin resistance (FDR=6,57e-03), and nervous system diseases (FDR=7,58e-05), among others (Table S4; Figure S6). These genes were differentially regulated in the brain; especially being up-regulated in the frontal cortex, anterior cingulate cortex, putamen, amygdala, nucleus accumbens, and hippocampus (Figure S7). A complete list of pathways enriched in agonistic and antagonistic loci can be found in Tables S5 and S6.

### Risk models for BMI and ΔBMI

Among the included variables in the Null models in the whole dataset, BMI_0_ contributed the most with 8.7% of the variance in ΔBMI_3_, AP treatment contributed 7.5% and sex 1.5%. BMI_0_ contributed 4.3% to the total variance of ΔBMI_12_, AP treatment 6.4%, and age 2.7% (Supplemental Information). If ΔBMI_3_ was included, it drastically improved the model for ΔBMI_12_, accounting for 28.6% of its variance (Figure 2).

**Figure 2.**
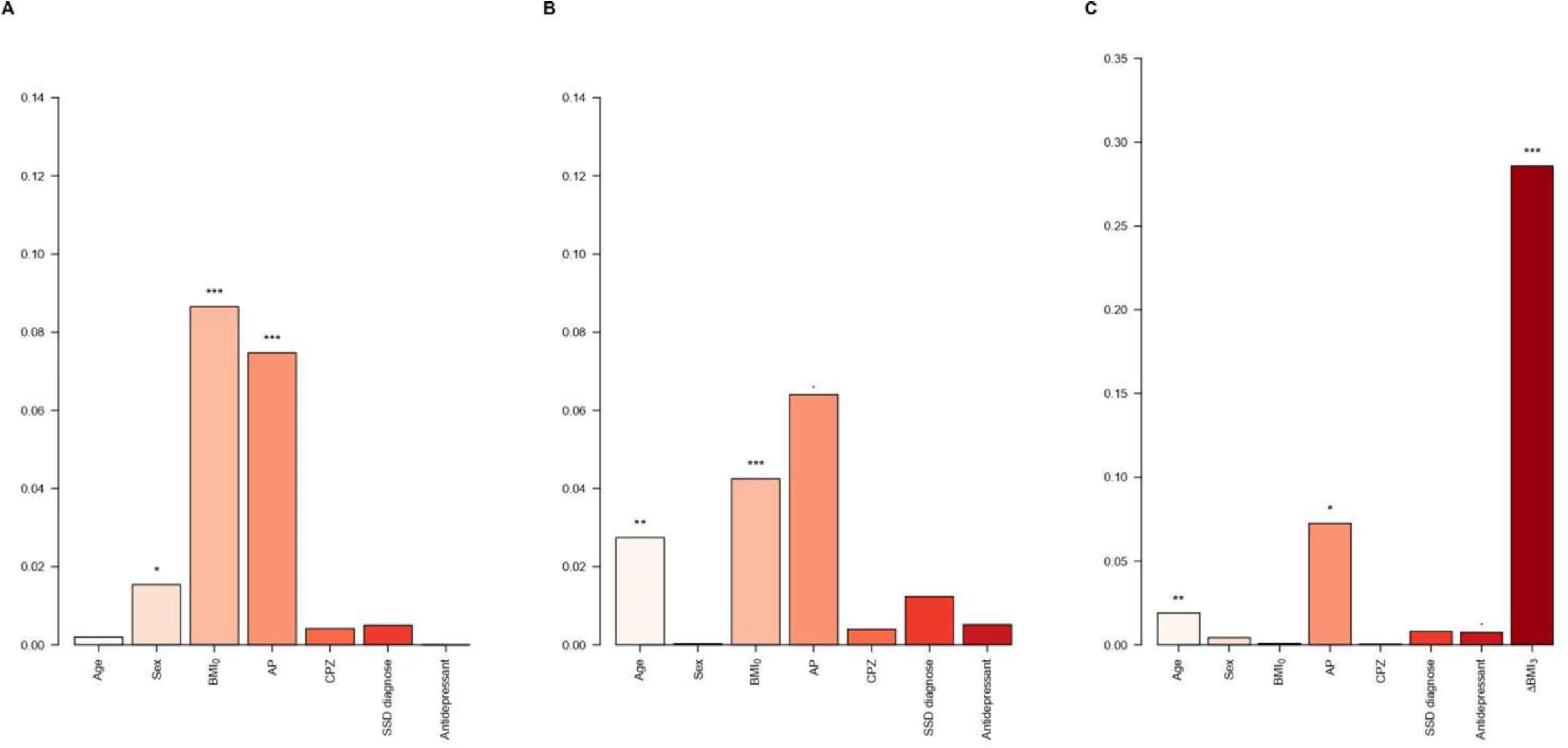
ΔBMI variance explained. Bar plots showing the variance explained by each covariate in the *Null models* for A) ΔBMI_3_, B) ΔBMI_12_, and C) ΔBMI_12_ including ΔBMI_3_ in the model. In the plots the variables for medication (either AP and CPZ) at 3m and 12m were merged. AP: Antipsychotic drug; CPZ: equivalent doses of chlorpromazine. *P value⍰<⍰0.05; **P value⍰<⍰0.01; ***P value<0.001.

PRS were calculated for BMI_0_, BMI_3_, and BMI_12_, the P_optimal_ in the Training set were established at 0.15, 0.15, and 0.08, respectively (Table S7). The *PRS models showed* an adjusted R^2^ (AdjR^2^) of 0.11 for BMI_0_ (P=0.01), 0.13 for BMI_3_ (P=0.01), and 0.21 for BMI_12_ (P=2.4e-03) in the Validation set (Table S8). In summary, the inclusion of PRS improved the Null models by 40% for BMI_0_, 32% for BMI_3_, and 55.5% for BMI_12_ (Figure 3). Next, we investigated the inclusion of PRS in predicting ΔBMI_3_ and ΔBMI_12_. The P_optimal_ thresholds in the *Training set* were estimated at 2.2e-03 and 0.01, for ΔBMI_3_ and ΔBMI_12_, respectively (Table S7). The *PRS models* showed AdjR2 of 0.19 for ΔBMI_3_ (P=1.3e-03) and 0.10 for ΔBMI_12_ (P=0.04, Table S9). The inclusion of PRS ostensibly improved the *Null models* by 50.7% in ΔBMI_12_ (P=0.02), but not ΔBMI_3_ (P=0.35). BMI_0_ is in part explained by the BMI PRS, so we decided to exclude it from the models. In this case, *PRS models* improved the *Null models* of both ΔBMI_12_ (177.3%, P=8.3e-03), and ΔBMI_3_ (21.2%, P=0.03, Figure 4).

**Figure 3.**
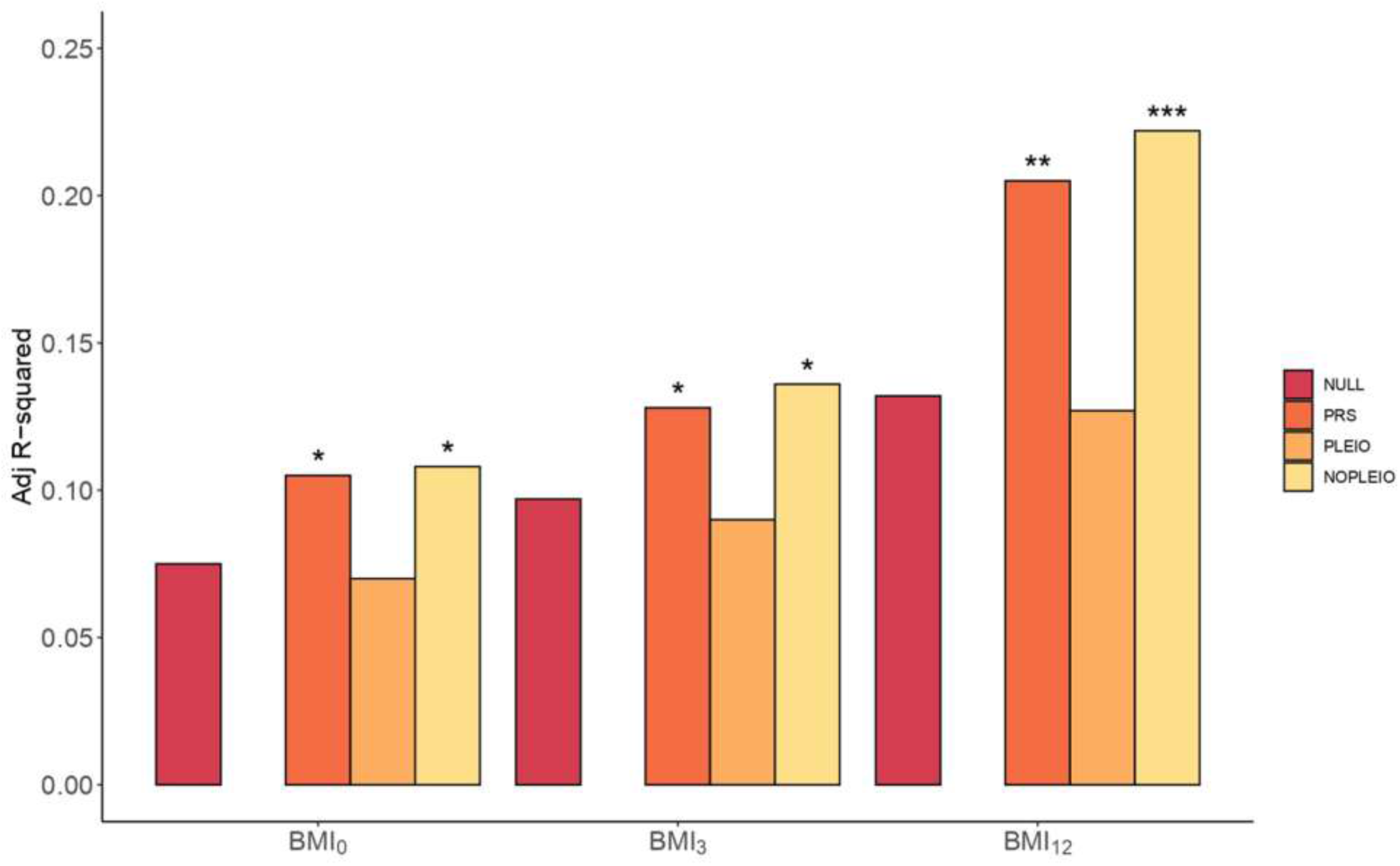
*Null* **vs** *PRS models* **in BMI**. Barplots showing the AdjR2 in BMI by the *Null model* and the *PRS model* computed using all SNPs (PRS), pleiotropic SNPs (PLEIO), and non-pleiotropic SNPs (NOPLEIO) from BMI GWAS. The barplot shows predictions of BMI_0_, BMI_3_ and BMI_12_, in the x-axis. Covariates included in the *Null model* were the first 10 PC, age, sex, AP drug prescribed, chlorpromazine equivalent doses, diagnose, and the presence or not of antidepressant treatment. Asterisks represent significantly improved models compared to the *Null models* (anova). *P value⍰<⍰0.05; **P value⍰<⍰0.01; ***P value<0.001.

**Figure 4.**
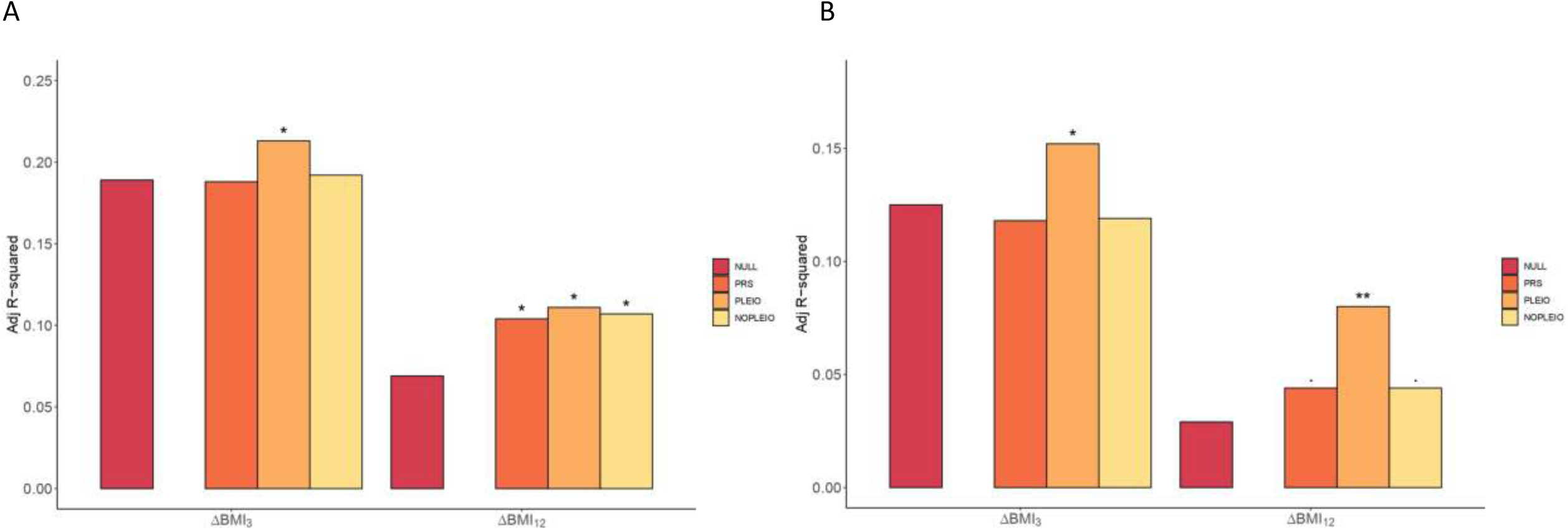
*Null* **vs** *PRS models* **in ΔBMI**. Barplots showing the AdjR^2^ (y-axis) in ΔBMI by the *Null model* (red), and *PRS model* computed using all SNPs (PRS), only pleiotropic SNPs (PLEIO), and non-pleiotropic SNPs (NOPLEIO) in the *Validation dataset* (n=157). A) Multiple linear models including BMI_0_ as a covariate in the model. B) Multiple linear models excluding BMI_0_ from the list of covariates. The rest of covariates included in the *Null model* were the 10 first PCs, AP treatment, chlorpromazine equivalent doses, age, sex, SSD diagnose, and being treated with antidepressant. Asterisks represent significantly improved *PRS models* compared to the *Null models* (anova). ·P value < 0.1; *P value⍰<⍰0.05; **P value⍰<⍰0.01; ***P value<0.001.

Next, we wanted to investigate whether loci shared between SCZ and BMI played any role in BMI and ΔBMI. So, we divided the genome-wide significant SNPs (P<5e-08) from BMI GWAS into pleiotropic and non-pleiotropic, using information from the conjunctional analysis, to obtain two BMI PRS based on pleiotropic and non-pleiotropic SNPs (Figure S8). The pleiotropic variants between SCZ and BMI (Table S10) played no role in determining BMI at any time (P=0.58, P=0.80, and P=0.58 for BMI_0_, BMI_3_, and BMI_12_, respectively), while variants coming from non-pleiotropic loci improved prediction in a similar way as all SNPs: by 44% for BMI_0_ (P=0.01), 40.2% for BMI_3_ (P=0.01), and by 68.2% for BMI_12_ (P=4.3e-04, Figure 3). In contrast, when evaluating ΔBMI, the *PRS model* obtained from the pleiotropic regions improved the ΔBMI_3_ model by a 12.7% (P=0.03) and the ΔBMI_12_ model by 60.9% (P=0.01, Figure 4). PRS obtained from non-pleiotropic regions was not relevant for predicting ΔBMI_3_ (P=0.25); however, it also significantly improved the *Null model* for ΔBMI_12_ by 55.1% (P=0.02). Finally, when the PRS was derived only from antagonistic loci, the risk models for both ΔBMIs outperformed the ones obtained with agonistic loci, being similar to those obtained with the inclusion of pleiotropic PRS (Figure S9).

If BMI_0_ was not considered in the models, inclusion of PRS from the pleiotropic loci increased by 21.6% (P=0.03) the ΔBMI_3_ model, and by 175.9% the ΔBMI_12_ (P=8.3e-03, Figure 4C). In contrast, non-pleiotropic loci did not improve any of the *Null models* without BMI_0_ (P=0.76 and P=0.1 for ΔBMI_3_ and ΔBMI_12_, respectively).

## Discussion

Understanding the genetic vulnerability associated with increased BMI carries the potential to predict its risk prior to treatment initiation and implement personalized risk-based treatments. In this study, we demonstrate for the first time, the replicable effects of BMI PRS in predicting ΔBMI in FEP.

In our study, BMI_0_ was inversely associated with ΔBMI_3_ and ΔBMI_12_, indicating that individuals with lower BMI_0_ were more likely to have higher ΔBMI during the first year of treatment (44). Remarkably, ΔBMI_12_ was strongly predicted by ΔBMI_3_ (explaining 30% of its variance), underscoring the importance of the first weeks of treatment on the outcome of obesity associated with psychosis, emphasizing the idea that clinicians should put the focus on early weeks to prevent long-term weight gain (45). In the analyses of patients who did not switch AP, our results are in accordance with previous studies in which a greater gain is observed in patients treated with atypical AP (46,47). However, the effects varied greatly within medications and interactions with underlying individual characteristics and genetic factors become relevant(48).

We confirm and extend with further data the shared genetic architecture between BMI and SCZ, involving pathways such as DNA damage, DNA methylation, insulin resistance, and dopaminergic and glutamatergic synapses in weight gain (26). These molecular pathways become promising mechanisms for understanding weight gain in FEP. Specific DNA methylation signatures have been widely described to play a role in obesity and weight loss in humans (49–51). There is also evidence pointing to the relevance of the glutamatergic system as a promising strategy to treat obesity (52). Also, weight gain associated with clozapine may be linked to antagonism of the histaminergic H1 receptors, increasing the risk for insulin resistance and type 2 diabetes (53).

Here, we demonstrate the role of genetics underlying both BMI and ΔBMI in FEP patients. The inclusion of the BMI PRS ostensibly improved the absolute BMI prediction. This is not surprising, as the role of genetics in BMI in the general population is already known (29), yet this is the first time it is also validated in FEP individuals. Notably, the inclusion of BMI genetic scores also improved the prediction of medium term ΔBMI. And strikingly, the inclusion of the PRS of pleiotropic regions between SCZ and BMI improved the risk model of ΔBMI_12_ and ΔBMI_3_. This observation was made stronger when BMI_0_ was not accounted for in the models. Our results also indicate a greater role for antagonistic variants in predicting ΔBMI. However, it could be simply a matter of statistical power as more SNPs were recovered in this category. In contrast, the genetic architecture of BMI not shared with SCZ played a pivotal role in controlling BMI. All these results highlight the importance of including patient genetic information to improve the prediction of weight gain in FEP patients.

Our study has one main strength; it is based on two independent longitudinal and prospective cohorts of well-characterized drug-naïve FEP patients. However, our work has some limitations; first, we could not control for well-known factors that contribute to BMI changes, such as diet and physical activity (54,55). Secondly, the interpretation of the results could also be skewed by the fact that FEP patients often switch AP treatment during follow-up, and are sometimes treated with a secondary AP and other psychiatric medications, all of them with a possible impact on BMI (56). Moreover, there may have been unrecorded extra changes in the AP prescriptions between the study timepoints. We have tried to reduce this problem by including current and past AP treatment in the models. Noteworthy, the two cohorts do not have the same distributions of AP treatments. Although this may imply a bias when comparing results between AP drugs, it also reinforces the role of genetics in ΔBMI given its replicability even in sets of individuals with different treatments. Future research with larger cohorts and including populations of non-European ancestry could provide even more information on genetic vulnerability to ΔBMI.

In summary, our findings highlight that genetics is an important factor in determining weight gain in patients with FEP, suggesting its inclusion in the clinical routine to identify individuals at higher risk, and to optimize individualized prevention programs and management to improve their quality of life. In turn, our findings pave the way to address the prediction of comorbid trajectories in other diseases using a similar approach.

## Data Availability

All data produced in the present study are available upon reasonable request to the authors

## Acknowledgements

This work was supported by the Catalan Agency of Research and Universities (AGAUR, 2017SGR-00444, PI: EV). GM is supported by Instituto de Salud Carlos III (PI18/00514 and PI21/00612). The Santander (PAFIP) cohort was funded by the following grants: Instituto de Salud Carlos III (FIS00/3095, PI020499, PI050427, PI060507), Plan Nacional de Drogas Research (2005-Orden sco/3246/2004), SENY Fundatio Research (2005-0308007), Fundacion Marques de Valdecilla (A/02/07, API07/011) and MINECO/FEDER (SAF2016-76046-R, SAF2013-46292-R). JV-B is supported by funding from Instituto de Investigación Valdecilla (INT/A21/10, INT/A20/04). AN is supported by funding from AEI-PGC2018-BI00 (FEDER/UE)(MINECO/FEDER, UE), “Unidad de Excelencia María de Maeztu”, funded by the AEI (CEX2018-000792-M), Secretaria d’Universitats i Recerca and CERCA Programme of the Departament d’Economia i Coneixement de la Generalitat de Catalunya (GRC 2017 SGR 880).

## Conflicts of Interest

BC-F has received honoraria (advisory board and educational lectures) and travel expenses from Takeda, Menarini, Angelini, Teva, Otsuka, Lundbeck, and Johnson & Johnson. He has also received unrestricted research grants from Lundbeck. JV-B has received honoraria for his participation as a consultant and/or as a speaker at educational events from Janssen-Cilag and Lundbeck. The rest of authors report no biomedical financial interests or potential conflicts of interest.

## Notes

### Author Declarations

The Clinical Research Ethics Committee of Cantabria approved the research protocol.

