## Supplemental Information for "Leveraging genetics to improve Body Mass Index increase prediction in the first-episode of psychosis"

**Supplemental Information Content**

Supplemental Methods	page 2

Supplemental Results	page 2

Supplemental References	page 5

Supplemental Figures	page 6

**Supplemental Methods**

**Quality control**

Quality control was performed with PLINK to exclude: single-nucleotide polymorphisms (SNPs) with a minor allele frequency (MAF) <0.05, genotyping failure >2%, a Hardy-Weinberg equilibrium p-value <10e-06; individuals with inconsistent sex and with heterozygosity levels 3 standard deviations away from the mean; and related individuals (the relative with the lower call rate). In addition, samples showing evidence of population stratification were identified by principal component (PC) analysis and excluded. The top 10 PC scores were saved for further analysis. SNP imputation was conducted with the Michigan Imputation Server^2^ against the full European reference panel. The imputed SNPs underwent another round of quality control, SNPs with MAF < 0.001, a Hardy-Weinberg equilibrium p-value <10e-06, --geno 0.01, --mind 0.01, and an imputation information score <0.9 were excluded. At the end, 6,910,431 SNPs were included in the *Training set* and 6,552,380 SNPs in the *Validation set*.

**Identification of Independent SNPs by pleiofdr**

Independent genomic loci were identified merging any physically overlapping lead SNPs (LD blocks < 250 kb apart), as described previously. Significant SNPs were identified as SNPs with a conjFDR < 0.05 and at LD r^2^ < 0.6 with each other^3^. Then, the borders of the genomic loci were defined by identifying all SNPs in LD (r^2^ ≧ 0.1) with one of the independent significant SNPs in the locus. The region containing all these candidate SNPs was defined as a single independent genomic locus and the most significant SNP within the region was selected as the lead SNP. Finally, we kept as pleiotropic all the significant independent SNPs identified by the software at conjFDR<0.05. The directional effects of the loci shared between SCZ and BMI measures were evaluated by comparing their z-scores. All p-values were adjusted for standard genomic control (GC) and Z-scores were adjusted for sample overlap between GWAS, using intergenic SNPs as implemented in *pleioFDR*. The European populations from the 1KGP project were used as the reference panel for the computation of the LD structure between SNPs.

**Supplemental Results**

**Differences among AP drugs in ∆BMI**

The effects of AP treatment on BMI were initially assessed in the 202 individuals that remained with the same AP drug during the period of the study. Although all APs were associated, on average, with a positive ∆BMI (Figure S4 and Supplementary Material), the AP drug was important for the extent of ∆BMI_3_ and ∆BMI_12_ (Kruskal-Wallis test; P=6.1e-04 and P=2e-03, respectively). Among them, olanzapine was the AP drug associated with the highest ∆BMI_3_ (12.1% mean increase) and ∆BMI_12_ (17.3% mean increase) and ziprasidone the one associated with the lowest outcomes (2.1% mean increase at ∆BMI_3_ and 5.0% at ∆BMI_12_).

These changes were consistently observed when including those patients that switched AP during the study. Altogether, 57 patients out of 224 (25.4%) in the *Training set* and 13 out of 157 (8.3%) in the *Validation* set changed their baseline AP medication to another one during the first 3 months. When comparing 3 and 12 months, 59 (29.2%) and 40 (29.0%) patients switched AP medication in the *Training* and *Validation* sets, respectively, being three and 18 patients the same that switched the AP between baseline and 3 months (Figure S2).

We compared ∆BMI across AP drugs including individuals that switched AP drugs during the study period. Significant differences in ∆BMI_3_ were observed according to the AP drug taken at baseline and at t=3m (P=3.7e-04 and P=3.9e-05, respectively). Those individuals treated with clozapine and paliperidone showed the largest ∆BMI_3_, while ziprasidone was associated with the lowest ∆BMI_3_ (Figure S10). After 12 months of treatment, patients at clozapine and paliperidone treatment at t=3m showed the greatest rise in BMI (32% and 38.2% ∆BMI_3_ mean increase, respectively; P=8.4e-03), while those treated with ziprasidone presented the lowest values (9% ∆BMI_12_ mean increase, Figure S11A). ∆BMI_12_ was also evaluated based on AP prescribed at t=12m, resulting also in significant statistical differences across AP drugs (P=1.8e-04). Paliperidone and clozapine were the AP with the highest ∆BMI_12_ (26.8% and 23.2%) while ziprasidone and quetiapine were the AP associated with lower ∆BMI_12_ (7.7% and 10.1%, Figure S11B). Finally, the use of antidepressant treatment during the first 3 months did not show significant differences in ∆BMI_3_ and ∆BMI_12_ (P=0.71 and P=0.44, respectively), nor the specific diagnosis (P=0.70 and P=0.05, respectively).

**Enrichments in agonistic and antagonistic SNPs**

SNPs with agonistic pleiotropy between SCZ and BMI (n=2,686) were mapped to 293 genes with ANNOVAR. Among them, the list was enriched in GO processes such as axon guidance (FDR=1,44e-03), but also pathways such as MAPK signaling pathway (FDR=4,72e-04), signal transduction (FDR=2,10e-05), and insulin resistance (FDR=2,11e-02, eTable 6). Among the antagonistic SNPs (6,864 SNPs corresponding to 631 genes) enriched categories included glutamatergic synapse (FDR=2,75e-10), positive regulation of neuron projection development (FDR=1,96e-04), cAMP signaling pathway (FDR=1,07e-06), axon guidance (FDR=9,89e-05), nicotinic acetylcholine receptor signaling pathway (FDR=5,42e-05), HDACs deacetylate histones (FDR=2,52e-16), and DNA methylation (FDR=1,21e-15). Also these genes were enriched in nervous system diseases (FDR=5,15e-03), among others (a complete list of enriched pathways can be found in eTable 5).

**Null models**

We first evaluated the *Null models* of BMI at each timepoint in the whole dataset. Variables included in the *Null models* were sex, age, diagnosis, being under antidepressant treatment, AP drug prescribed at the time of evaluation, and dose of chlorpromazine equivalents. Among the included variables, male sex was significantly associated with an increased BMI_0_, BMI_3_, and BMI_12_; age was significantly associated with BMI_0_ and BMI_3_; AP treatment was significantly associated with BMI_3_ and BMI_12_, being on haloperidol, olanzapine, risperidone and quetiapine (only in BMI_12_) drugs associated with higher BMI. Neither the diagnosis, the equivalent doses of chlorpromazine, nor the treatment with antidepressants were significant at any time (eTable 11). Secondly, we evaluated the null models for ∆BMIs. The multiple linear model for ∆BMI_3_ showed an adjusted R^2^ (adjR^2^) of 0.19 (P= 4.24e-11). Variables that were significant for ∆BMI_3_ were: BMI_0_ (P=5.9e-09), and being treated with olanzapine (P=4.2e-04) that significantly raised ∆BMI_3_, while female sex (P=0.01) and being treated with ziprasidone significantly reduced ∆BMI_3_ (P=9.4e-03). Age, diagnosis, being under antidepressant treatment, and dose of chlorpromazine equivalents did not show an effect on ∆BMI_3_ (eTable 12). Finally, the null model for ∆BMI_12_ showed an adjR^2^ of 0.17 (P=2.14e-09) and variables associated with higher ∆BMI_12_ were reduced age (P=1.8e-03), low BMI_0_ (P=1.1e-04), and treatment with paliperidone during the first 3 months (P=0.04, eTable 13).

**Supplemental Figures**

**Figure S1.** Boxplot of ∆BMI_3_ and ∆BMI_12_ in the whole dataset.

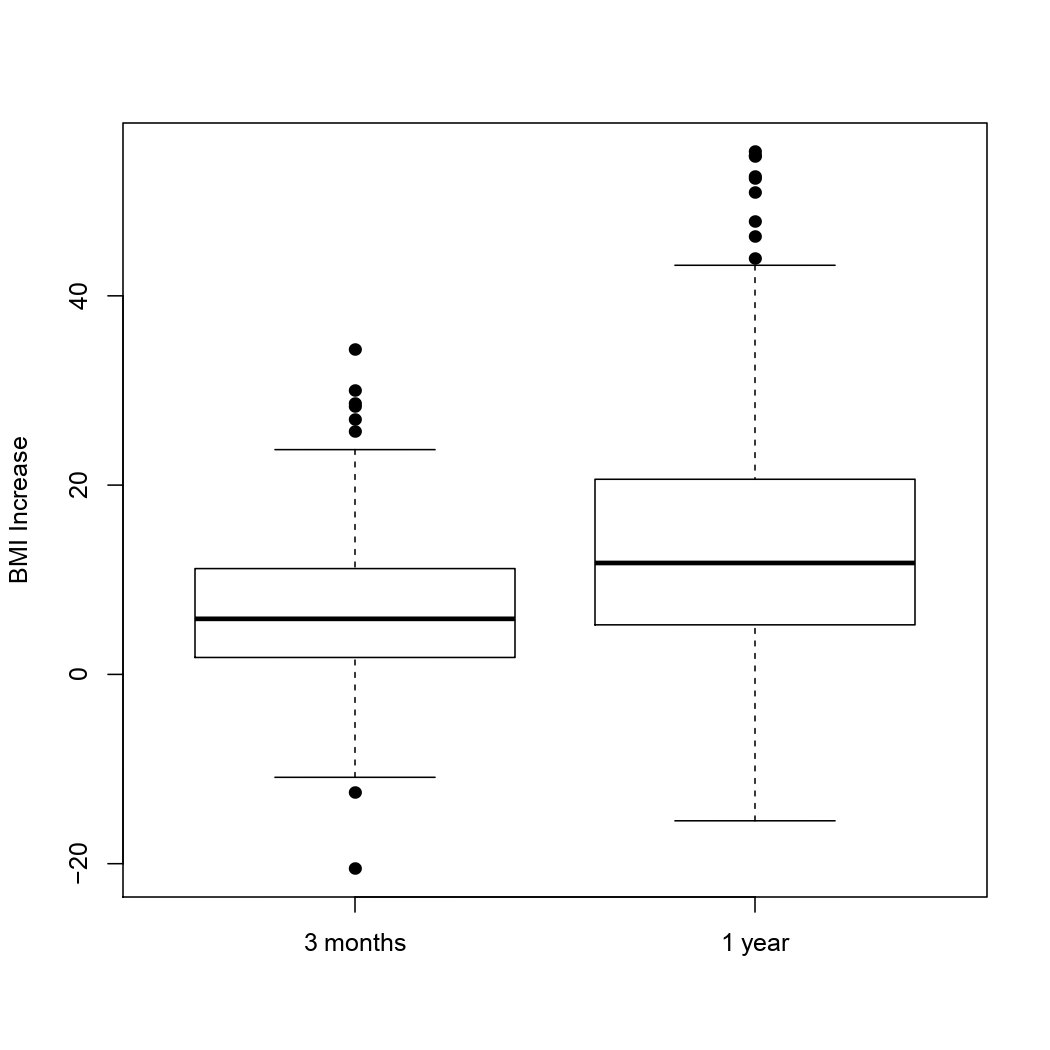

**Figure S2.** Graphic representation showing the individuals that discontinued the AP drug medication during the first year of the study. Both Validation and Training datasets are represented. Numbers in the boxes represent the patients that continued with the same AP treatment as baseline.

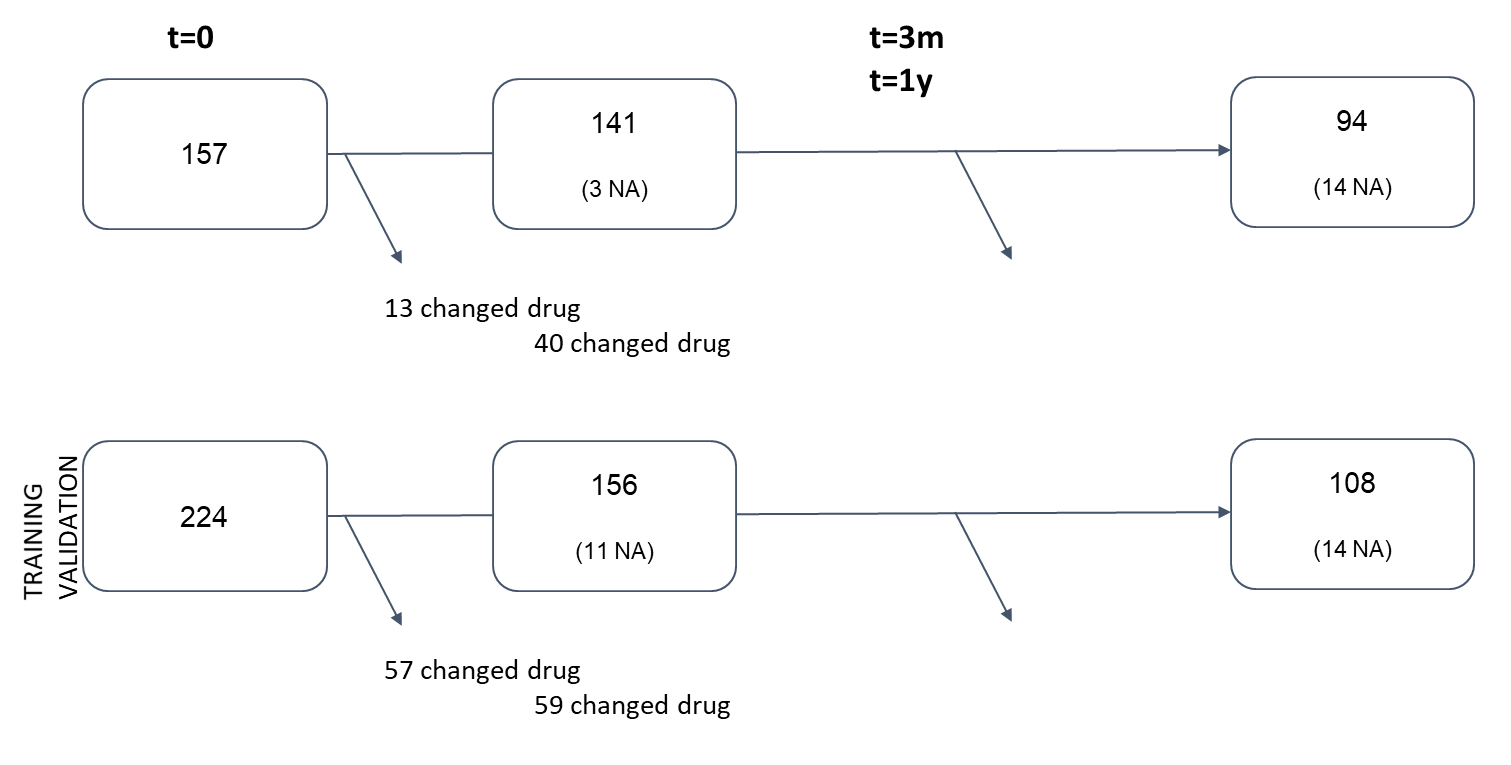

**Figure S3.** A) Correlation plot of BMI_0_, ∆BMI_3_, and ∆BMI_12_. Note that BMI_0_ was negatively correlated with all ∆BMI, while ∆BMI_3_ and ∆BMI_12_ were highly correlated among them. B) Scatterplot of Spearman's correlation of BMI_0_ (y-axis) and ∆BMI_3_ (x-axis) in 354 patients. C) Scatterplot of Spearman's correlation of BMI_0_ (y-axis) and ∆BMI_12_ (x-axis) in 340 patients. The dotted red lines show the best fit. r= correlation estimate. P= p-value of the correlations.

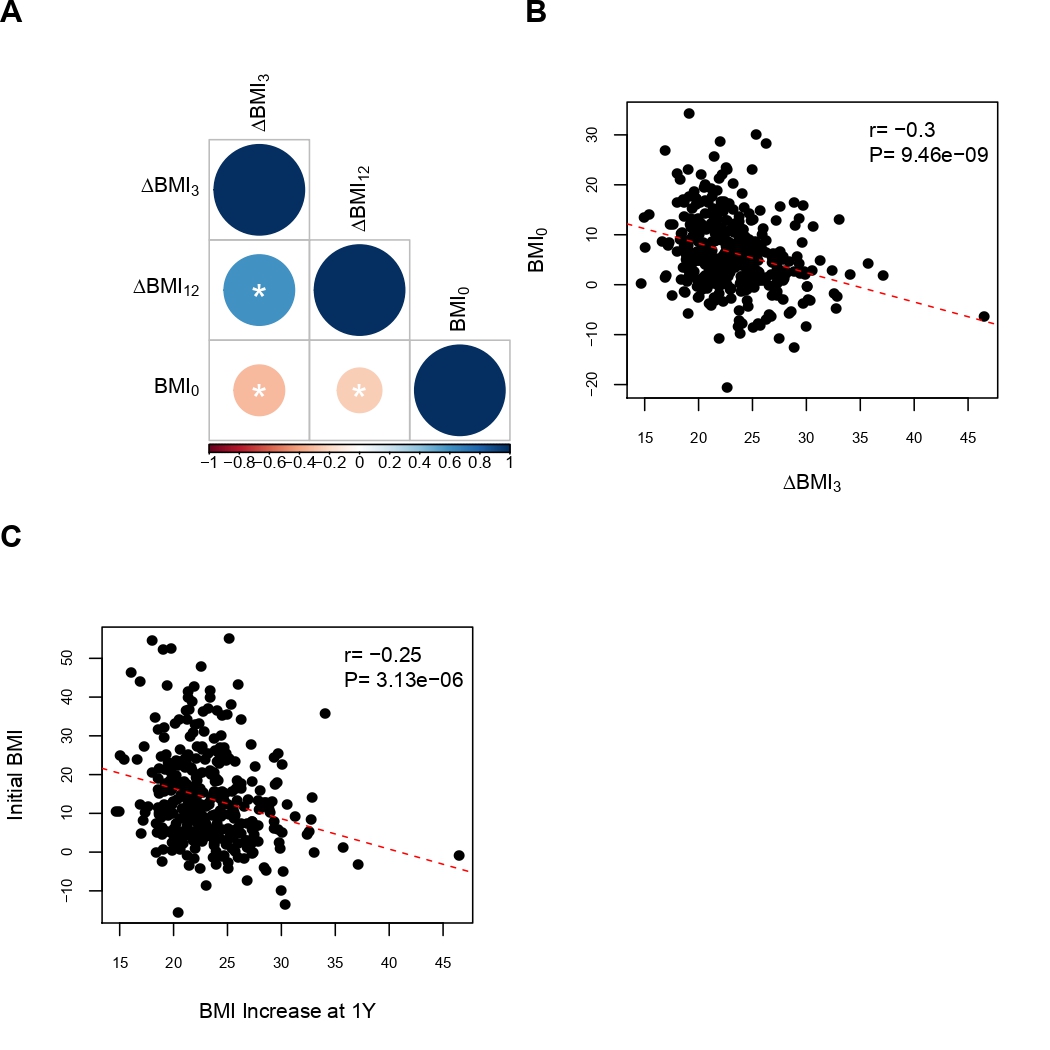

**Figure S4.** Boxplot of the A) ∆BMI_3_ and B) ∆BMI_12_, as a function of the AP drug (x-axes). Only the patients that did not switch AP during the follow-up have been considered (n=202).

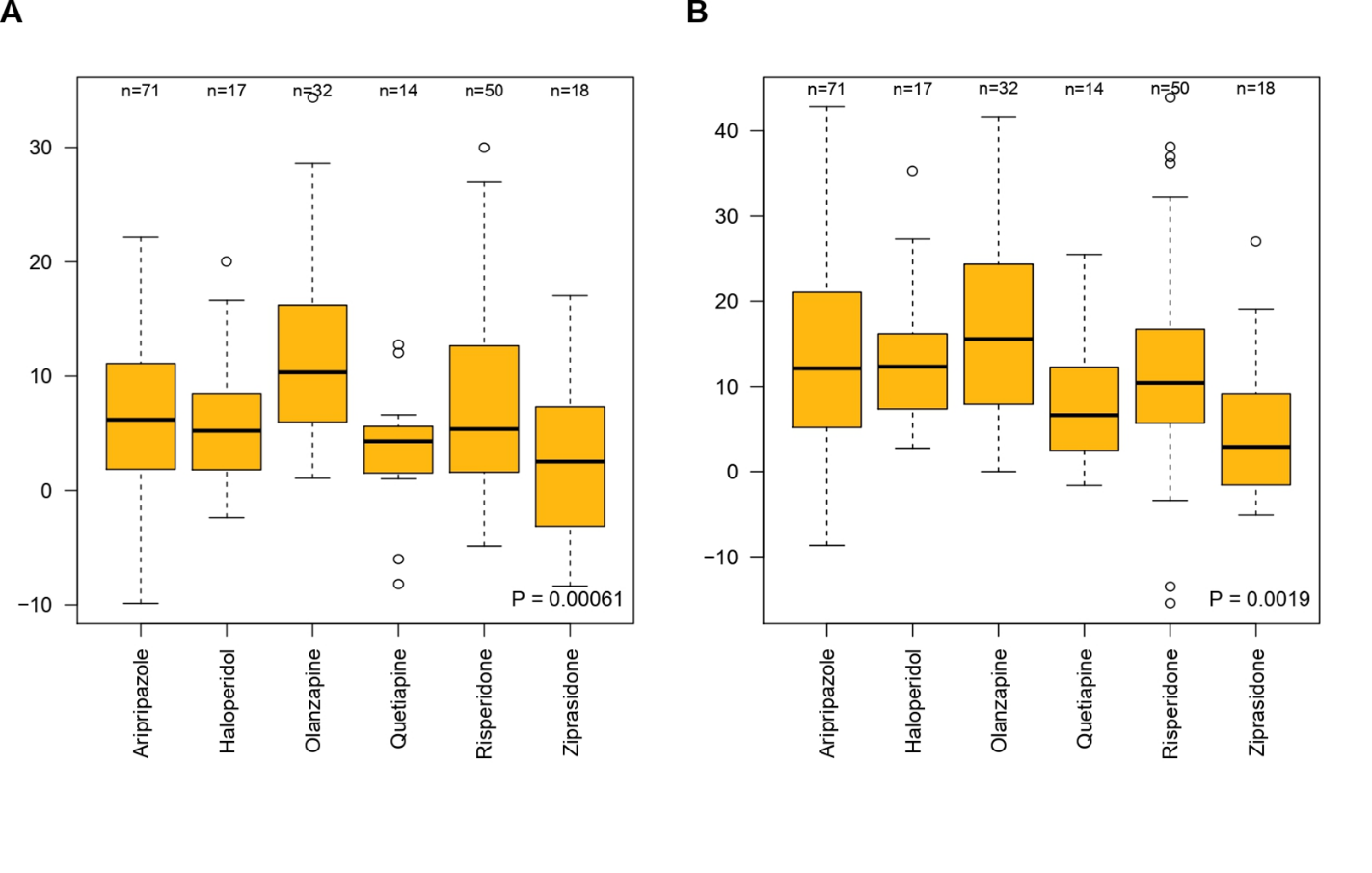

**Figure S5.** C) Ratio between antagonistic (negative) and agonistic (positive) pleiotropic variants across different conjFDR thresholds.

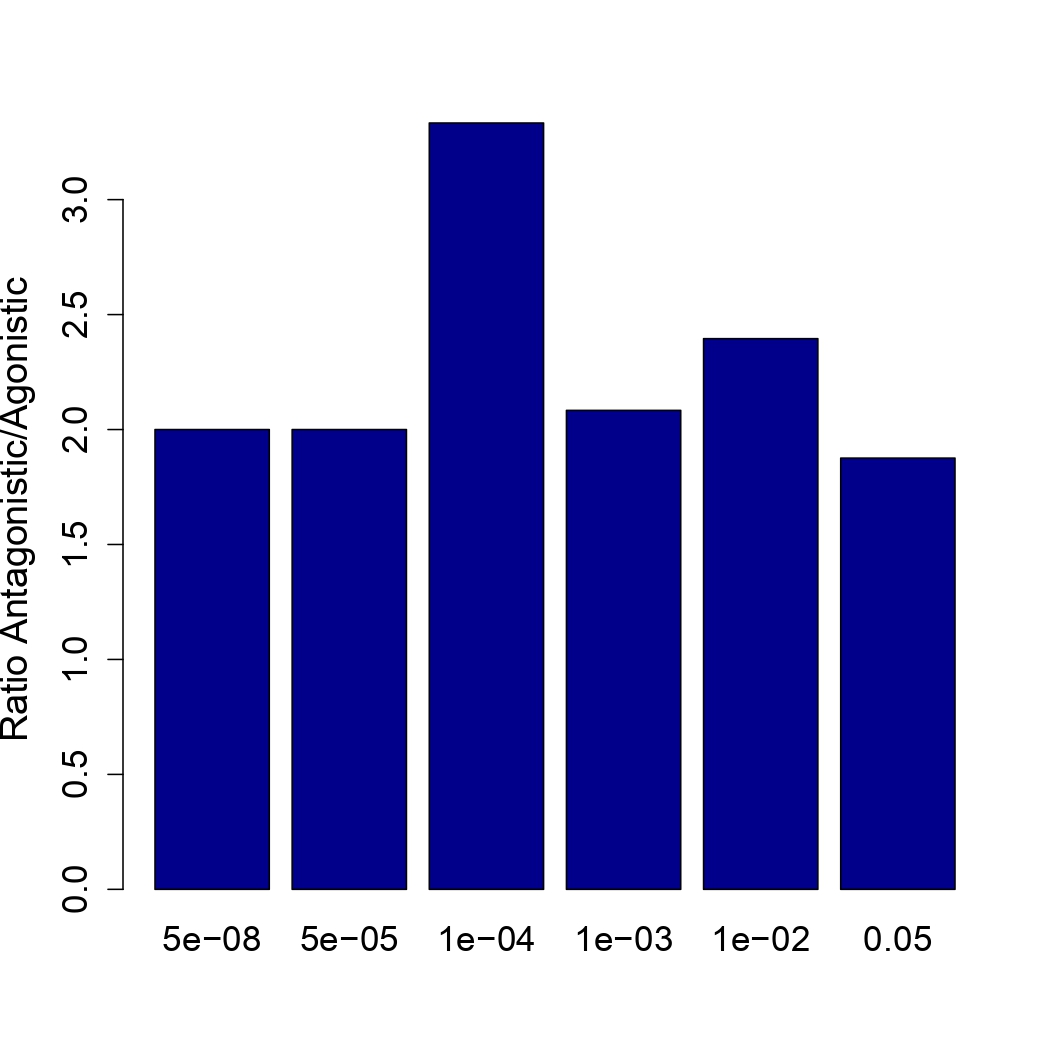

**Figure S6.** Bubble plot of enriched terms from KEGG pathways in A) all pleiotropic, B) only agonistic, and C) only antagonistic mapped genes. The color of the bar represents different clusters assigned by KOBAS-i. For each cluster, if there are more than 5 terms, top 5 with the highest enrich ratio will be displayed.

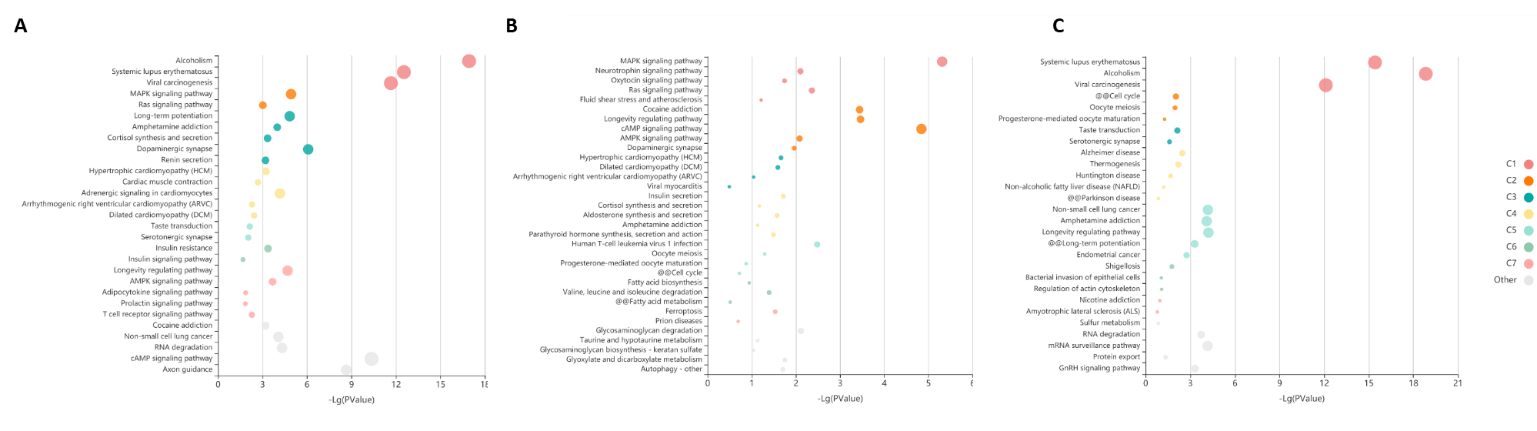

**Figure S7.** DEG in specific tissues for all SNPs at conjFDR<0.05, from GTEx v8 54 tissue types.

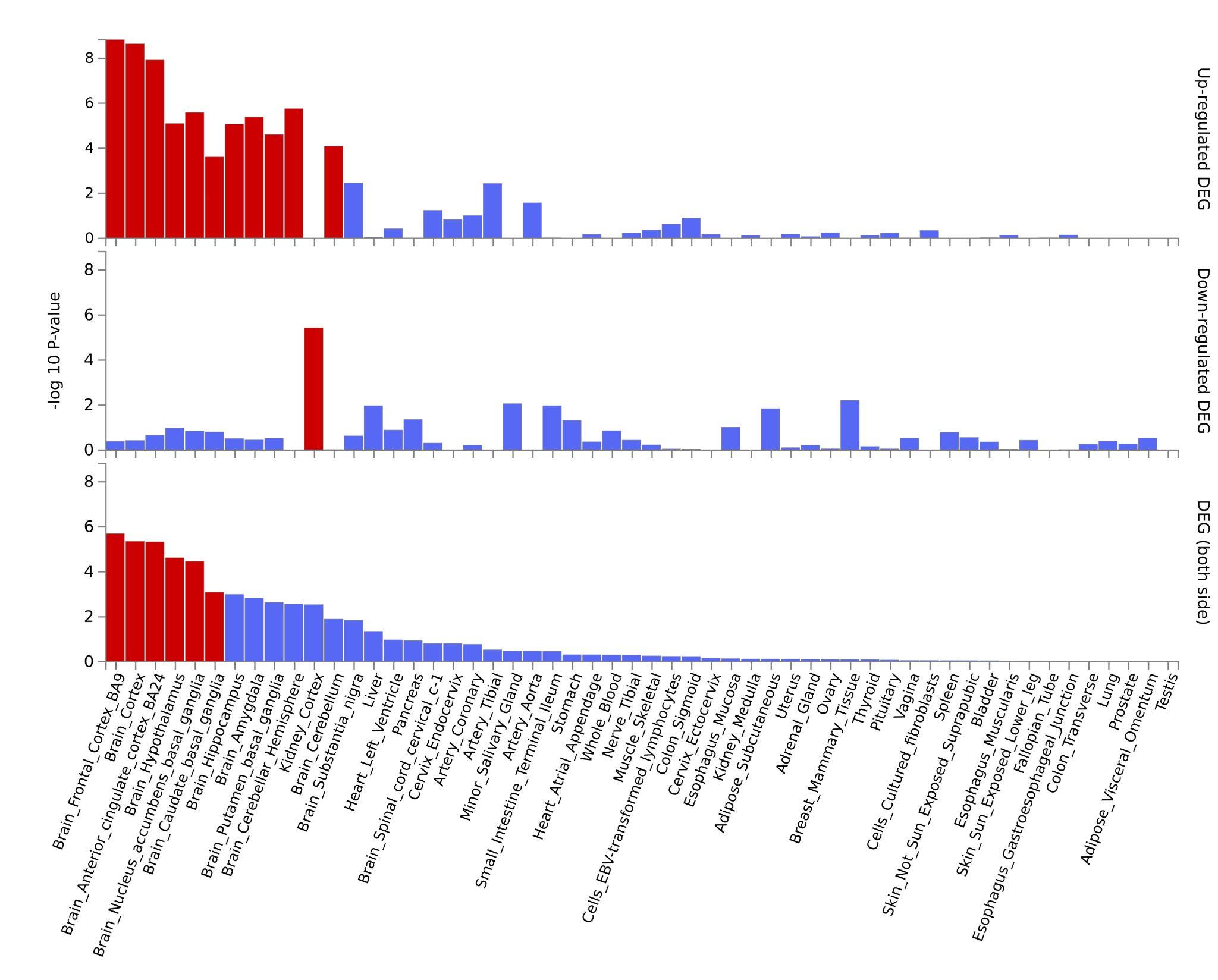

**Figure S8.** Manhattan plot of the Pulit et al. GWAS on BMI showing only the SNPs with pleiotropic information that were used for the PRS analyses. Green dots represent SNPs that were determined as pleiotropics betwen SCZ and BMI, while grey dots represent non-pleiotropic SNPs (the rest).

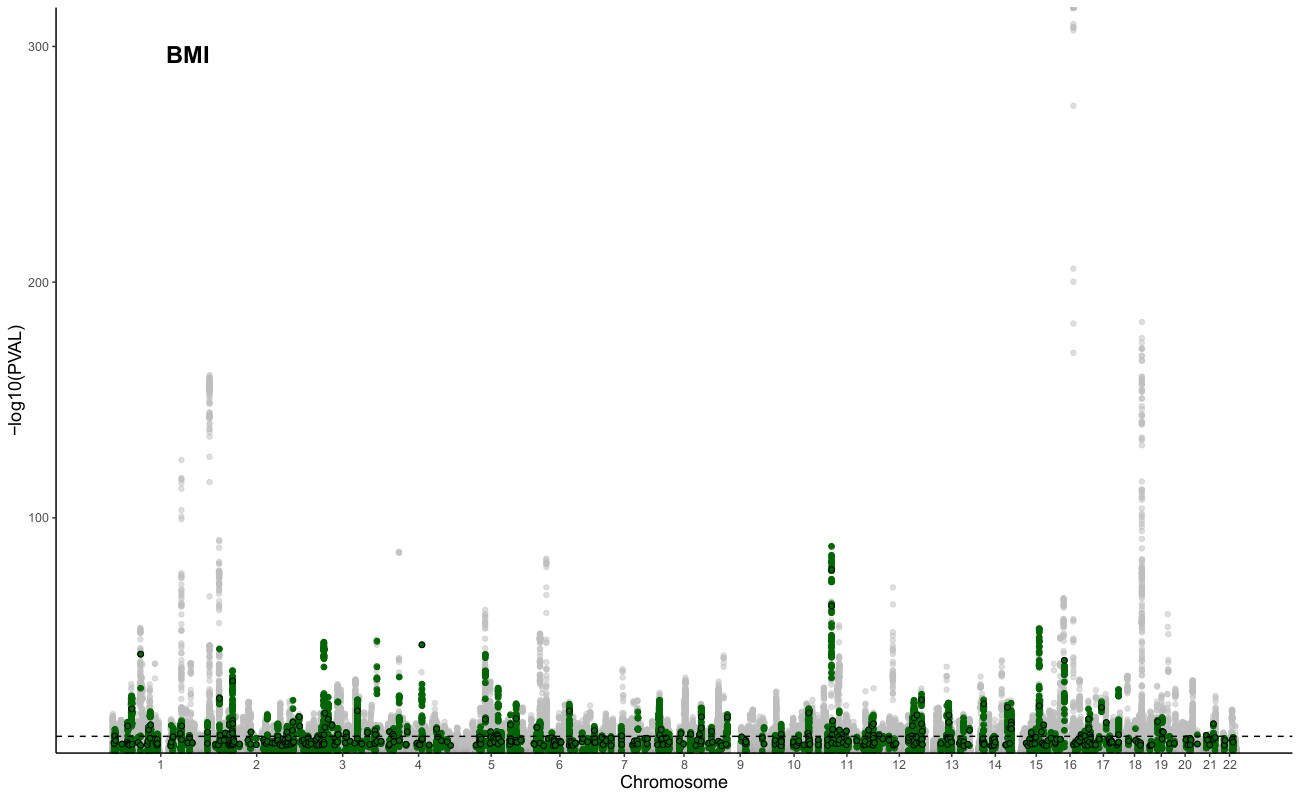

**Figure S9.** Bar plot showing the variance explained in total BMI at baseline (BASAL), including the Partitioned PRS of pleiotrpoic and non-pleiotropic regions. Asterisk refer to the significance in the independent ANOVA testing of the improvement from to the NULL model (in red).

* P-value < 0.05.

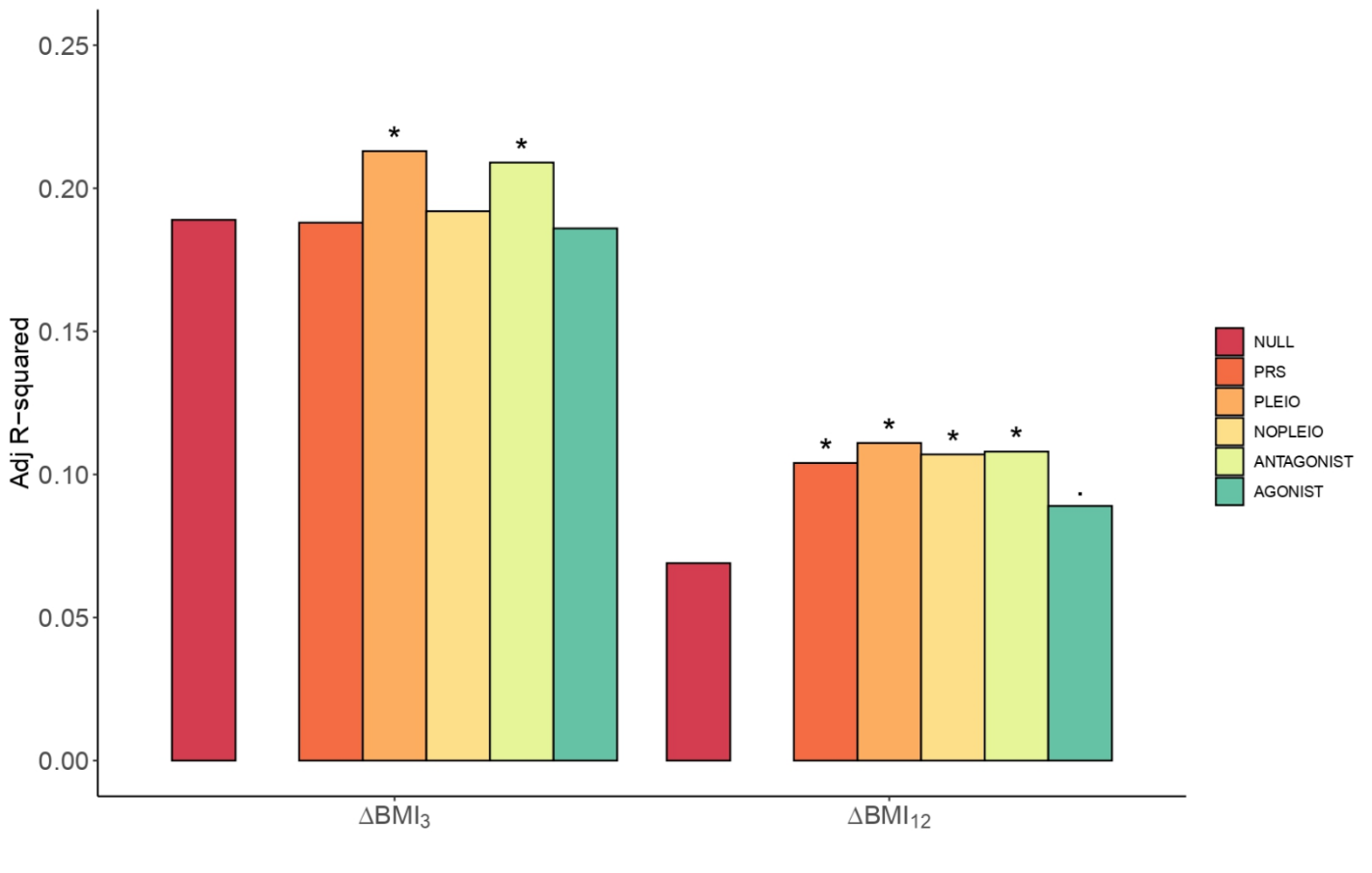

**Figure S10.** Boxplot of the ∆BMI_3_, as a function of the AP drug (x-axes) taken at A) baseline and B) 3 months after treatment initiation. Number of individuals in each group are shown in the top of the graphs and P-values obtained from the kruskal-Wallis test are shown at the bottom right of the graph.

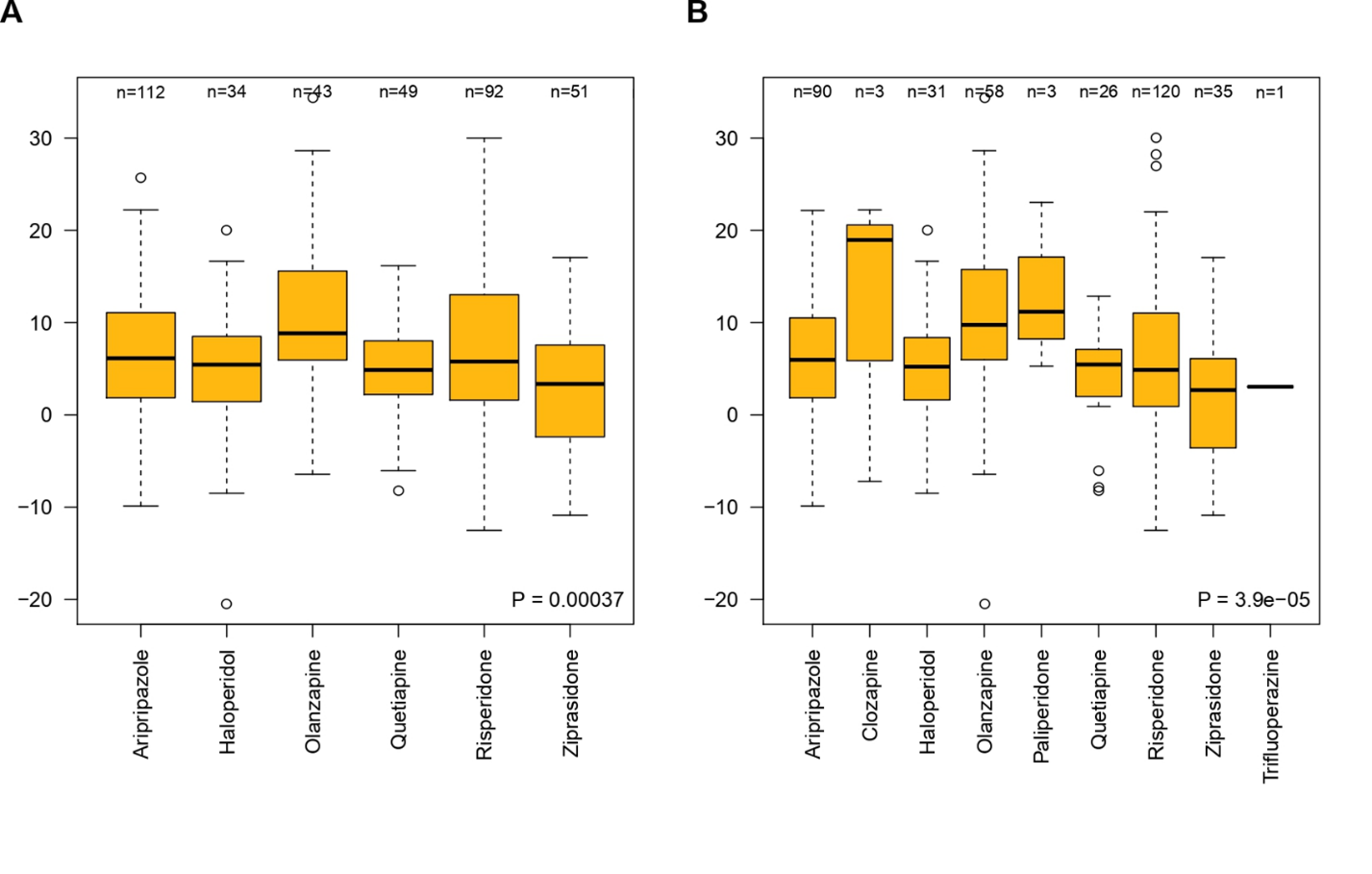

**Figure S11.** Boxplot of the ∆BMI_12_, as a function of the AP drug (x-axes) taken at A) 3 months and B) 1 year after treatment initiation. Number of individuals in each group are shown in the top of the graphs and P-values obtained from the Kruskal-Wallis test are shown at the bottom right of the graph.

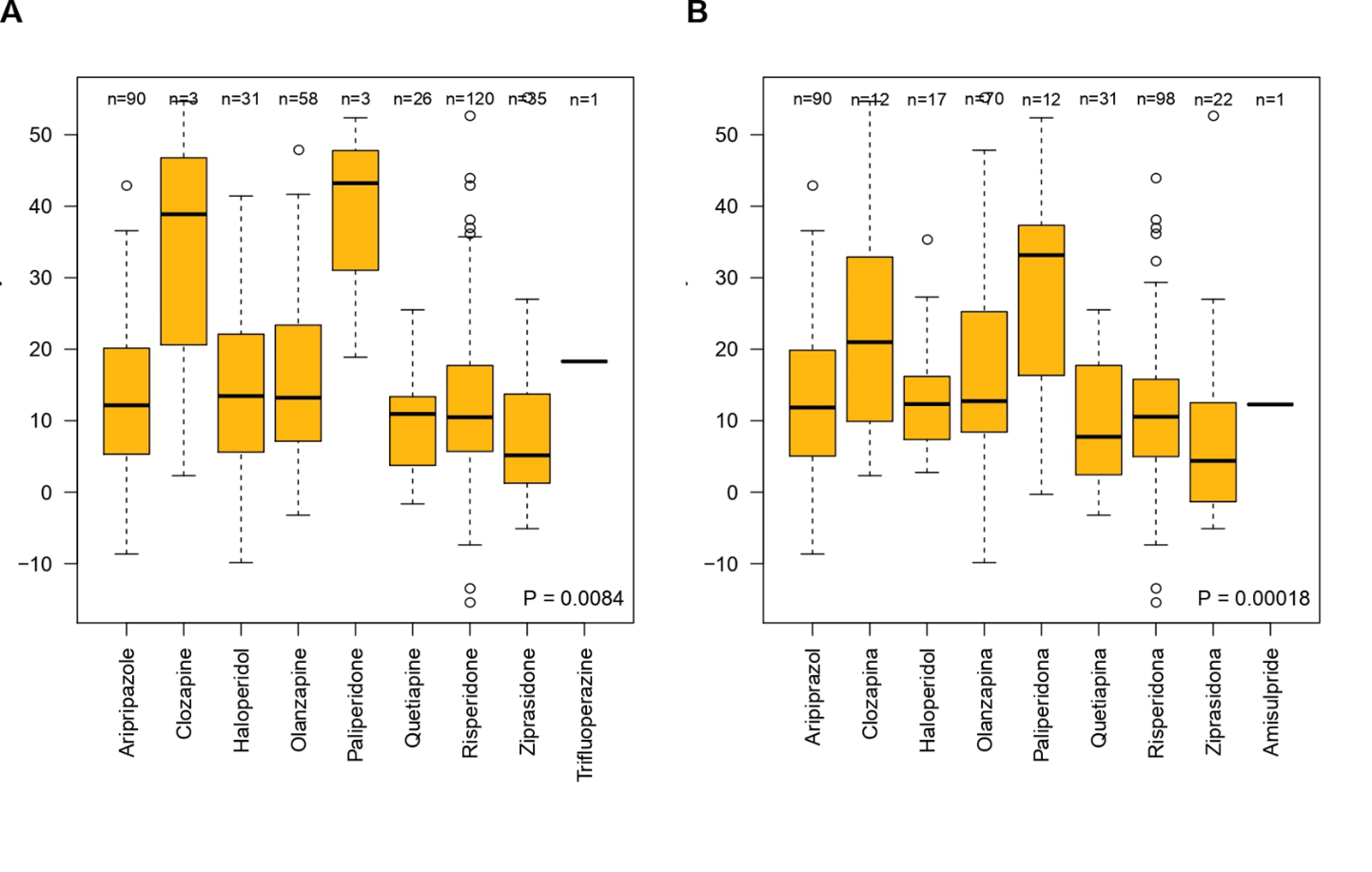
